# Longitudinal changes in IgG-type SARS-CoV-2 antibody titers after COVID-19 vaccination and, prominent increase in antibody titers when infected after vaccination

**DOI:** 10.1101/2023.02.04.23285414

**Authors:** Hiroshi Kusunoki, Michiko Ohkusa, Rie Iida, Ayumi Saito, Mikio Kawahara, Kazumi Ekawa, Nozomi Kato, Keita Yamasaki, Masaharu Motone, Hideo Shimizu

**Affiliations:** Department of Internal Medicine, Osaka Dental University, Hirakata, Osaka, Japan; Department of Laboratory Medicine, Osaka Dental University Hospital, Chuo-ku, Osaka, Japan; Department of Environmental and Preventive Medicine, Hyogo Medical University, Nishinomiya, Hyogo, Japan; Department of Health and Sports Sciences Graduate School of Medicine, Osaka University, Suita, Osaka, Japan; Faculty of Health Sciences, Osaka Dental University, Hirakata, Osaka, Japan

**Keywords:** COVID-19, SARS-CoV-2, vaccines, breakthrough infection, SARS-CoV-2 antibody

## Abstract

**[Objective]:** Severe acute respiratory syndrome coronavirus 2 (SARS-CoV-2) antibody titers level and duration of elevated levels are considered important indicators for confirming the efficacy of coronavirus disease 2019 (COVID–19) vaccines. The objective of this study was to demonstrate the changes in antibody titers after the second and third doses of the COVID–19 vaccine, and to determine the antibody titers in cases of spontaneous infection with SARS-CoV-2 after vaccination.

**[Materials and Methods]:** From June 2021 to February 2023, IgG-type SARS-CoV-2 antibody titers were measured in 127 participants, including 74 outpatients and 53 staffs, at the Osaka Dental University Hospital (65 males and 62 females, mean age 52.3 ± 19.1 years).

**[Results]:** Consistent with previous reports, the SARS-CoV-2 antibody titer decreased with time, not only after the second dose, but also after the third dose of the vaccine if there is no spontaneous COVID–19 infection. We also confirmed that the third booster vaccination was effective in increasing the antibody titer.

21 cases of natural infections were observed after administering two or more doses of the vaccine. 13 of these patients had post-infection antibody titers exceeding 40,000 AU/mL, and some cases continued to maintain antibody titers in the tens of thousands of AU/mL even after more than 6 months had passed since infection.

**[Conclusion]:** The rise in and duration of antibody titers against SARS-CoV-2 are considered important indicators for confirming the efficacy of novel COVID–19 vaccines. A longitudinal follow-up of antibody titers after vaccination in larger studies is warranted.

## Introduction

Severe acute respiratory syndrome coronavirus 2 (SARS-CoV-2) antibody titers level and duration of elevated levels are considered important indicators of the efficacy of novel coronavirus vaccines. In Japan, with the spread of coronavirus disease 2019 (COVID–19), many medical institutions offer the SARS-CoV-2 antibody titer test as an uninsured, self-paid test. However, there are few reports on antibody titer testing from such medical institutions. In Japan, SARS-CoV-2 mRNA vaccination began in February 2021, and more than 70% of the population had been vaccinated at least twice by the end of 2021. In Japan, the predominantly used mRNA vaccines for COVID–19 are the BNT162b2 mRNA vaccine (BioNTech and Pfizer) and the mRNA-1273 vaccine (Moderna and Takeda). It is generally known that antibody titers after SARS-CoV-2 vaccination decay over several months. In Japan, COVID–19 has spread rapidly due to the sixth wave of infection from the beginning of 2022, which was mainly caused by the Omicron strain.

Since the number of patients infected after the sixth wave was overwhelmingly larger than the cumulative number of those infected before the sixth wave, it can be assumed that breakthrough infection with COVID–19 after two or more vaccinations is very common. However, few reports have followed SARS-CoV-2 antibody titers over time after spontaneous infection following vaccination.

The Department of Internal Medicine, Osaka Dental University Hospital, has been measuring IgG-type SARS-CoV-2 antibody titers using the Architect SARS-CoV-2 IgG II Quant (Abbott Laboratory) in outpatients and medical staff since June, 2021.

In this article, we will show the relationship between the number of days after the second and third booster doses and antibody titers, as well as the evolution of antibody titers in cases of COVID-19 infection after more than two doses of the vaccine.

## Materials and methods

This was a single-center retrospective study. We conducted a longitudinal observational study involving outpatients and medical staff at the Osaka Dental University Hospital. The study protocols were approved by the ethics committee of Osaka Dental University Hospital (2022–10). Written informed consent was obtained from all participants. From June 2021 to February 2023, IgG-type SARS-CoV-2 antibody titers were measured in 127 participants, including 74 outpatients and 53 staff members at the Osaka Dental University Hospital (65 males and 62 females, mean age 52.3 ± 19.1 years). The titers of SARS-CoV-2 anti-receptor-binding domain IgG antibodies were measured in serum samples.

First, the correlation between antibody titer, subject age, and the number of days after the second dose of the vaccine was examined in 56 subjects (30 males and 26 females) with no history of COVID-19 infection, and the date of vaccination was clear. The same examination was also conducted on 23 subjects (12 males and 11 females) with no history of COVID-19 infection after the third dose of the vaccine, and the date of vaccination was clear. The correlation between pre-and post-vaccination antibody titers was examined in 13 subjects (six males and seven females) who had antibody titers measured before and after the third dose of the vaccination.

The post-infection antibody titers of subjects infected with COVID-19 after two or more doses of the COVID-19 vaccine were also examined.

## Serology assays

We used the Abbott Architect SARS-CoV-2 IgG II Quant (Abbott Laboratories, Illinois, USA) chemiluminescent microparticle immunoassay to detect IgG antibodies to the receptor-binding domain of the S1 subunit of the SARS-CoV-2 spike protein, according to the manufacturer’s instructions. The reportable measurement range of the assay is up to 40,000 AU/mL. IgG antibody titers >50 AU/mL (as the cut-off set by the manufacturer) were considered indicative of seropositivity.

## Statistical analysis

The results are expressed as mean ± standard deviation (SD). Pearson’s product-moment correlation coefficient was used to assess the associations between SARS-CoV-2 anti-receptor binding domain IgG antibody titers, age, and number of days after vaccination. For data analysis, the JMP 13.1 software was used for data analysis. Statistical significance was set at p < 0.05.

## Results

Antibody titers were measured in 56 subjects (30 males and 26 females) after the second dose of the vaccine. These subjects had no history of COVID-19 infection at the time of antibody titer measurement. A tendency for antibody titers to decrease was observed. The mean antibody titer was 4807.7 ± 6472.6 AU/mL (Figure 1).

**Figure 1.**
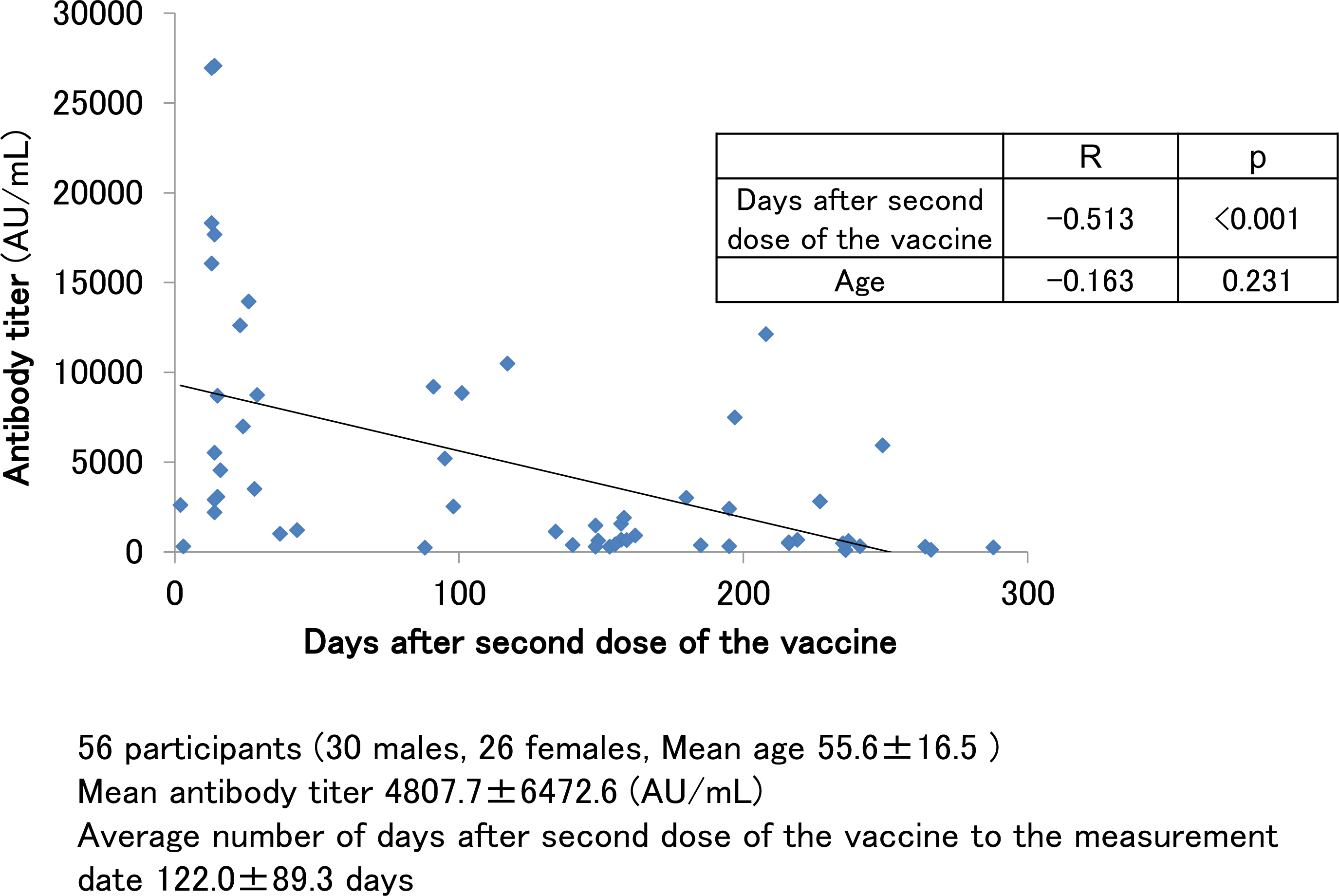
SARS-CoV-2 antibody titers in participants with no history of COVID–19 infection and had antibody measured after the second dose of the vaccine.

The antibody titer was negatively correlated with the number of days after the second vaccine dose (p < 0.001). It has previously been reported that antibody titers after vaccination tend to decrease with increasing age (1–4). A mild correlation between age and antibody titer was observed in our cohort of subjects, but the association was not statistically significant (p = 0.231).

Figure 2 shows the longitudinal changes in antibody levels in eight patients (four males and four females) with several antibody titer follow-ups after the second dose of the vaccine. These eight patients had no history of COVID-19 infection during that time. The first antibody titer was measured within 1 month of the second dose of the vaccination, but the titers varied greatly among individuals, ranging from several thousand to several tens of thousands AU/mL. The titers decreased by half to one-third in 3 months, and by 6 months, the titers decreased to a few hundred AU/mL.

**Figure 2.**
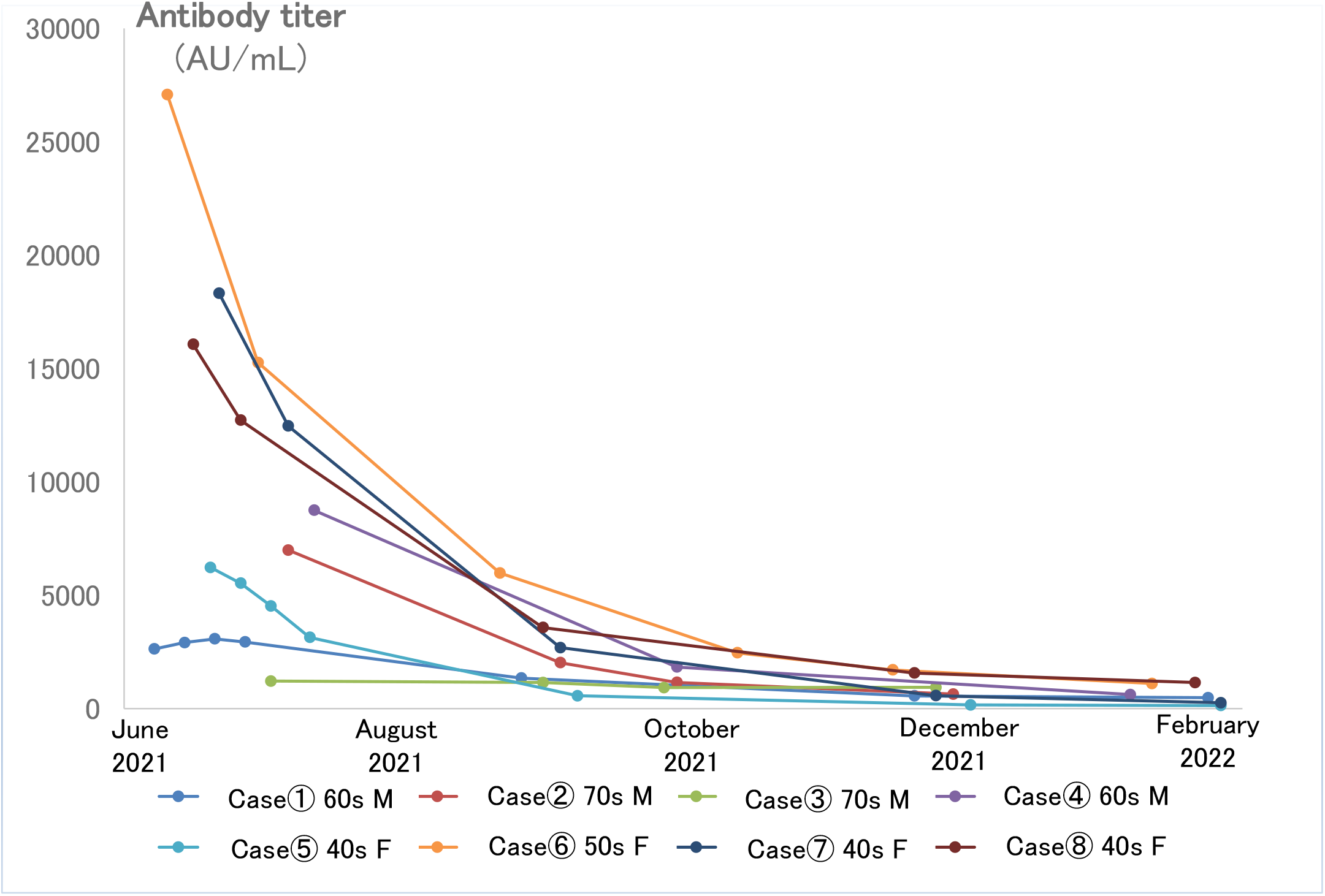
Longitudinal changes of SARS-CoV-2 antibodies in 8 cases with multiple follow-ups after the second dose of the vaccine.

For these eight cases, we compared the maximum antibody titer and the minimum antibody titer. The antibody titer decreased remarkably before the third dose of the vaccine. When the difference between the maximum and minimum titers was divided by the number of days between the two samplings, it was found that the higher the maximum value, the higher the rate of decrease in the antibody titer per day (Table 1).

**Table 1.** Eight cases with multiple follow-ups after the second dose of vaccine

| Case | Days from 2nd dose of the vaccine<br>to maximum antibody titer | Maximum antibody titer<br>(AU/mL) | Minimum antibody titer<br>(AU/mL) | Follow number of days | Rate of decrease in antibody titer<br>(AU/mL/day) |
| --- | --- | --- | --- | --- | --- |
| Case① 60s M | 21 | 3073.8 | 475.5 | 230 | 11.3 |
| Case② 70s M | 24 | 6988.8 | 635.4 | 154 | 41.3 |
| Case③ 80s M | 65 | 1211.7 | 936.1 | 154 | 1.8 |
| Case④ 60s M | 29 | 8745.5 | 616.8 | 189 | 43.0 |
| Case⑤ 40s F | 7 | 6221.9 | 132.2 | 234 | 26.0 |
| Case⑥ 50s F | 14 | 27079.9 | 1101.9 | 228 | 113.9 |
| Case⑦ 40s F | 13 | 18316.8 | 252.3 | 166 | 108.8 |
| Case⑧ 40s F | 13 | 16064.3 | 1144 | 232 | 64.3 |

Antibody titers were measured in 23 subjects (12 males and 11 females) with no history of COVID-19 infection at the time of antibody titer measurement after the third booster dose of the vaccine (Figure 3A). The average antibody titer after the third dose of the vaccine was 12617.5 ± 7256.2 AU/mL, which was certainly boosted from that after the second dose of the vaccine. The number of days between the third dose of the vaccine and the antibody titer measurement date also showed a negative correlation with the antibody titer, as did the titer after the second dose of the vaccine (p = 0.039). As in the case after the second dose of the vaccine, there may be a slight correlation with age, but this was not significant in our cohort of subjects (p = 0.212).

**Figure 3.**
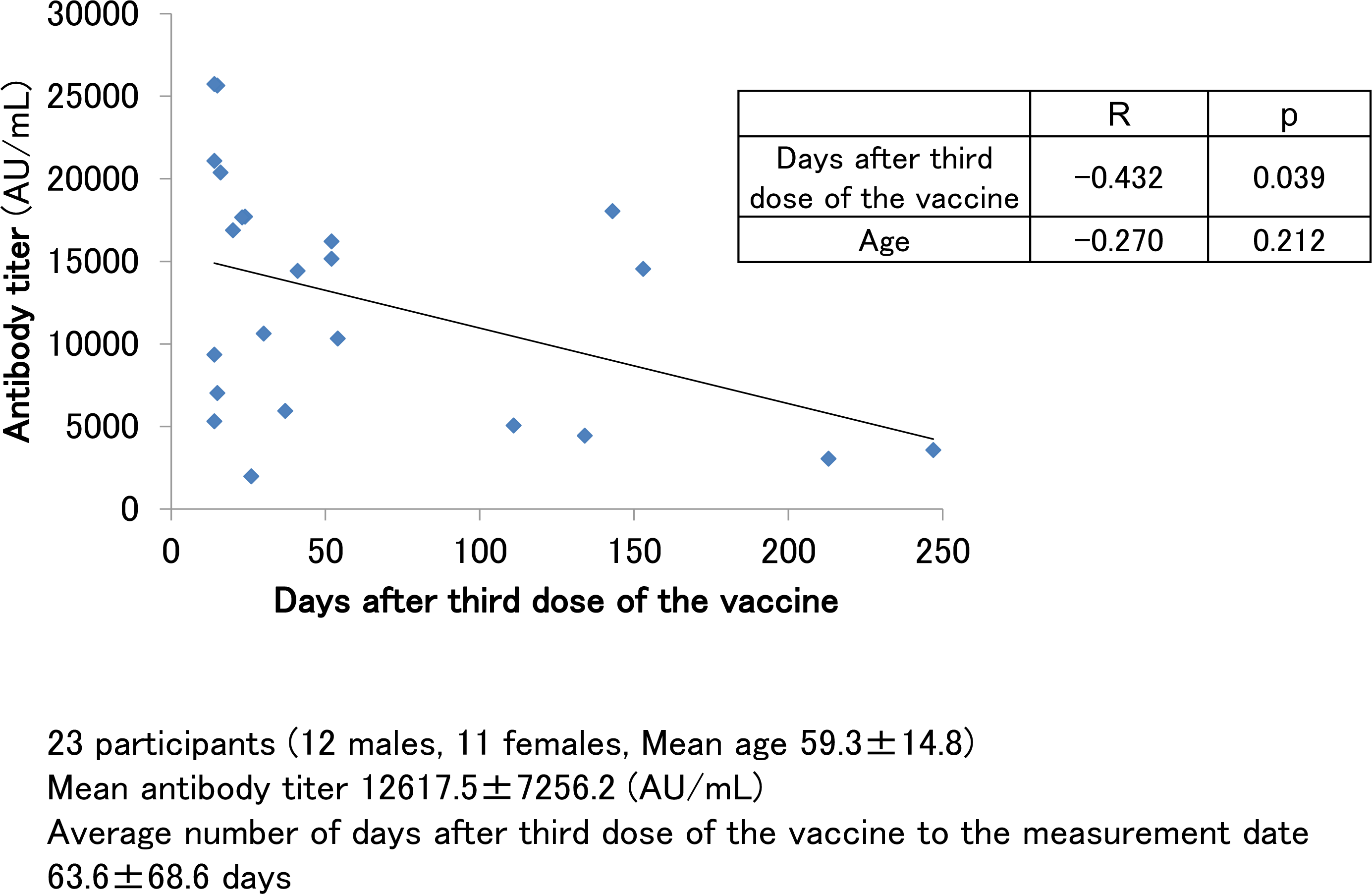
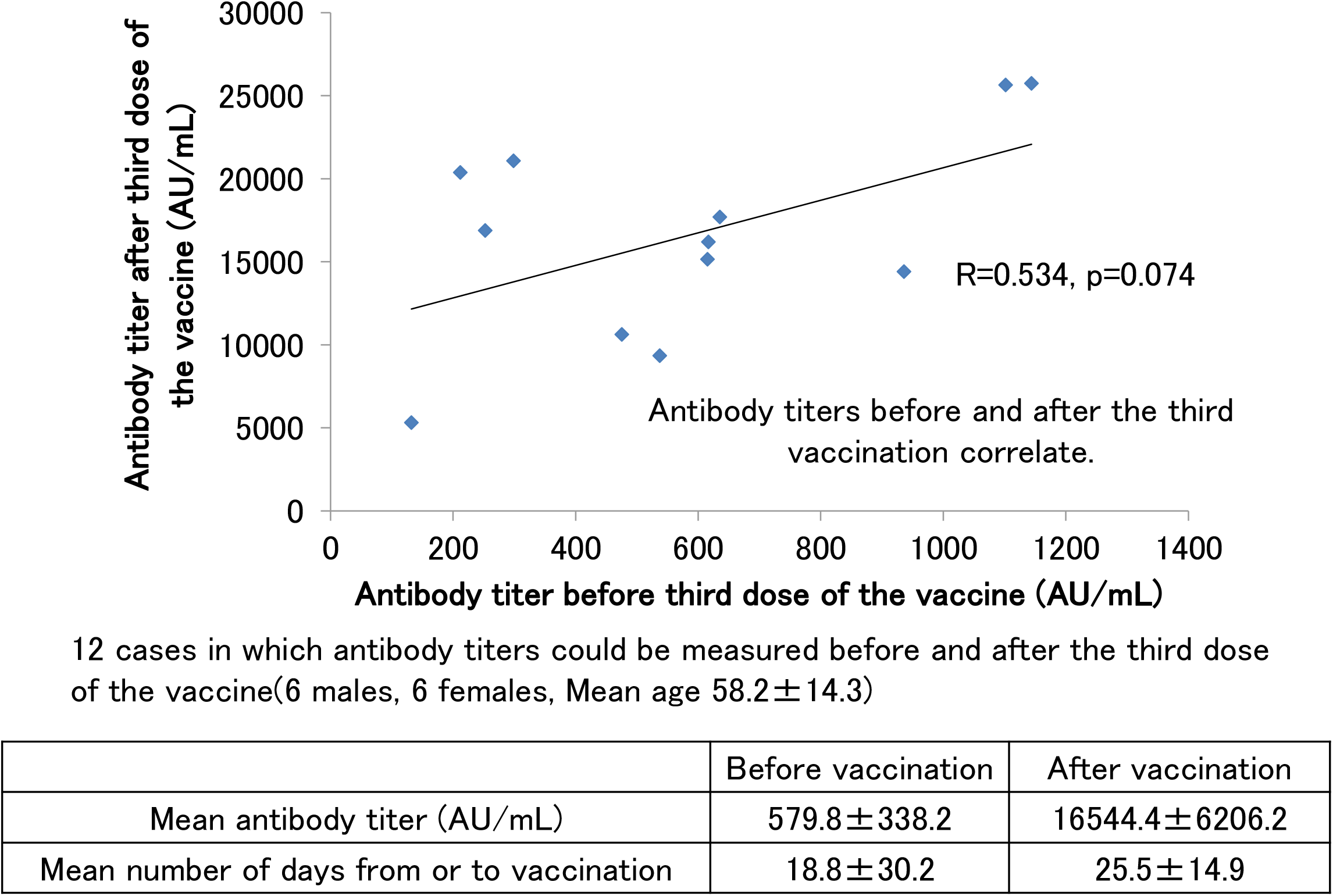
**A**. SARS-CoV-2 antibody titers in participants with no history of COVID-19 infection and had antibody measured after the third dose of the vaccine. **B**. Correlation of SARS-CoV-2 antibody titers before and after the third dose of the vaccine.

Figure 3B shows the scattered plotting of the antibody titers before and after the third booster dose of the vaccine for the 12 subjects (6 males and 6 females) from whom serum samples could be obtained before and after the third booster dose of the vaccine. This number of subjects did not result in a significant correlation (p = 0.074), but the correlation coefficient was above 0.5, indicating a significant correlation. It was suggested that a positive correlation exists between antibody titers before and after the third booster dose. We also found that the average antibody titer before the third booster dose of the vaccine was in the range of several hundred AU/mL, but after the third booster dose, many of the subjects had titers above 10,000 AU/mL.

Next, the cases of patients with boosted antibody titer after two doses of the vaccine and who developed COVID–19 infection are shown (Figure 4A). The antibody titer of a 40s male before the first dose of the vaccine in July 2021 was negative, and he had no episode of suspected COVID–19 infection up till that time. After the second dose of vaccine, his antibody titer jumped to nearly 27,000 AU/mL, and then decreased to about 4,000 AU/mL in approximately six months. When the antibody titers were measured in April 2022, he was considering the third dose of vaccine, but his titers increased to over 40,000 AU/mL, which was well above the limit of measurement. In other words, it is thought that this participant was spontaneously infected with COVID–19 somewhere from February to March 2022, but PCR testing was not conducted, and the participant was not counted as a COVID–19 infected patient. Even nine months after the suspected COVID–19 infection, the antibody titer remained above 4,0000 AU/mL.

**Figure 4.**
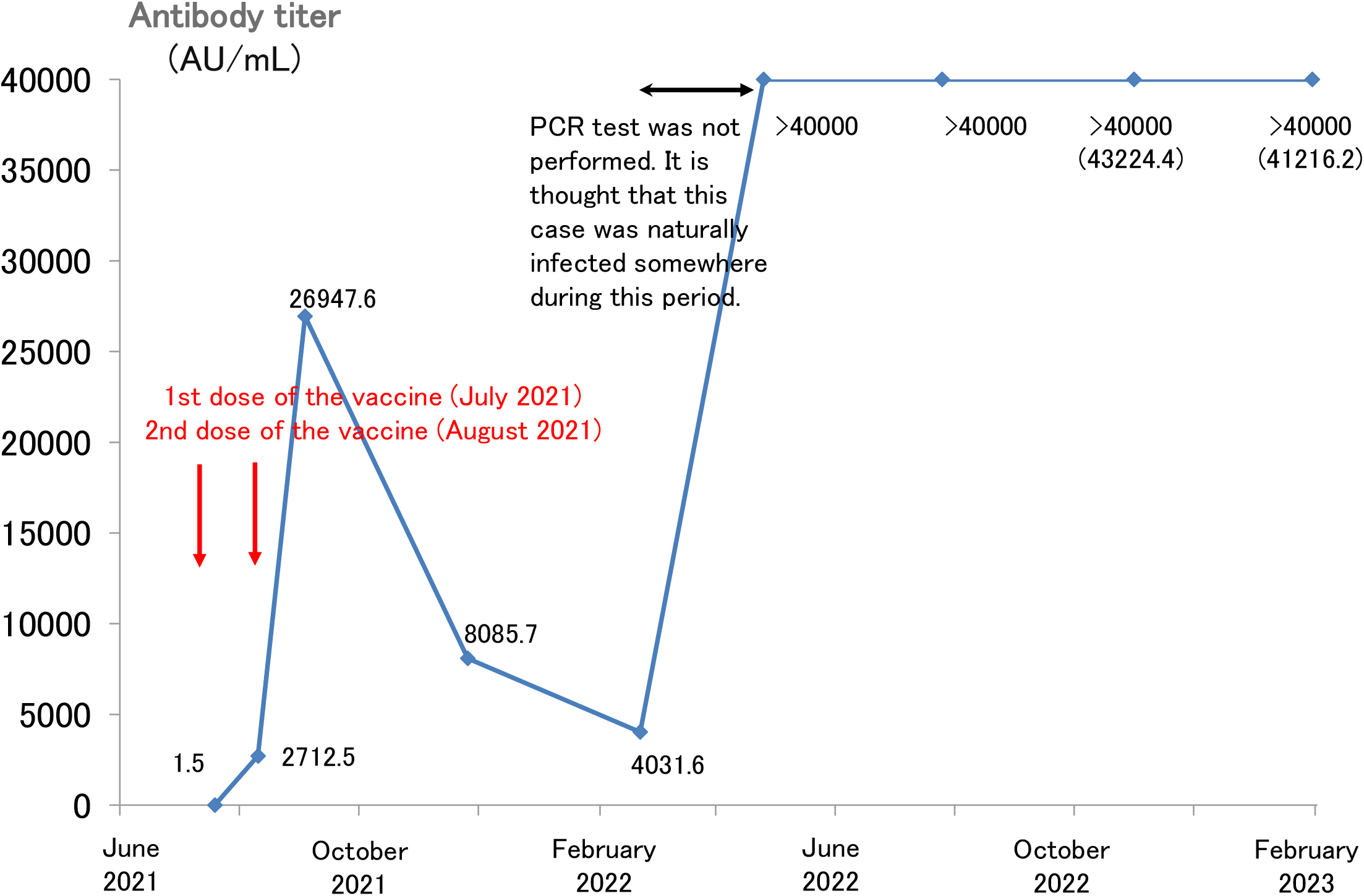
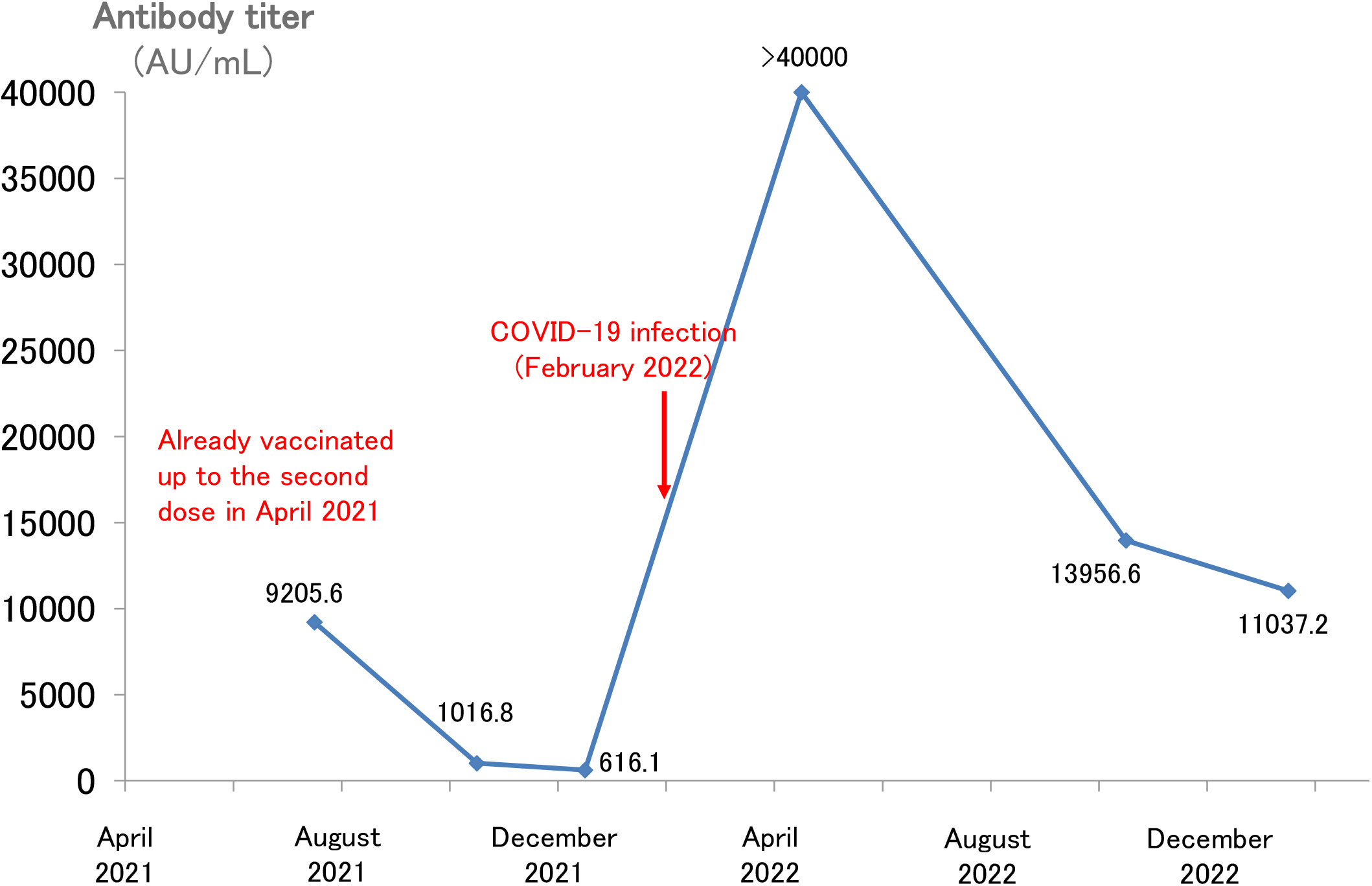
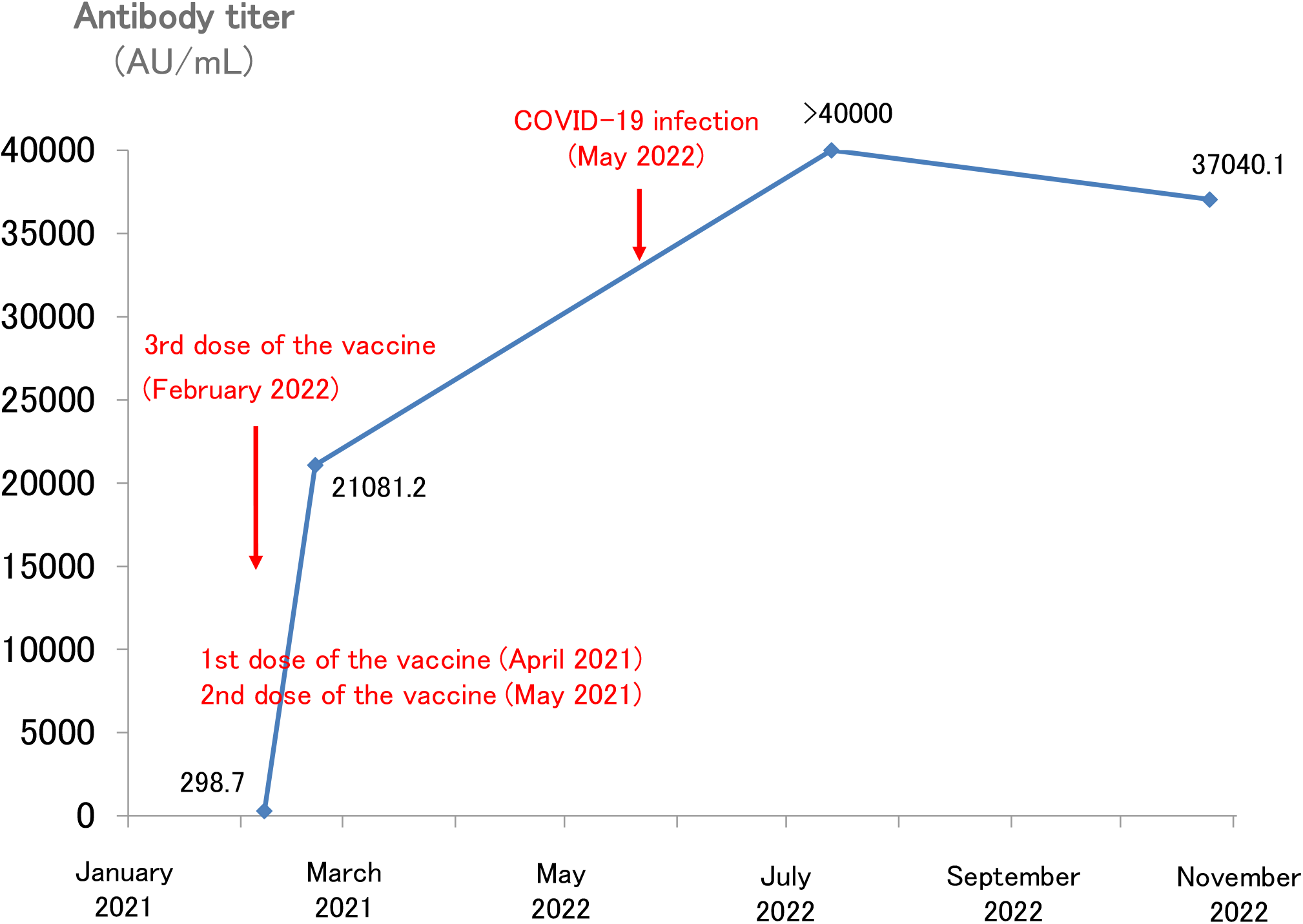
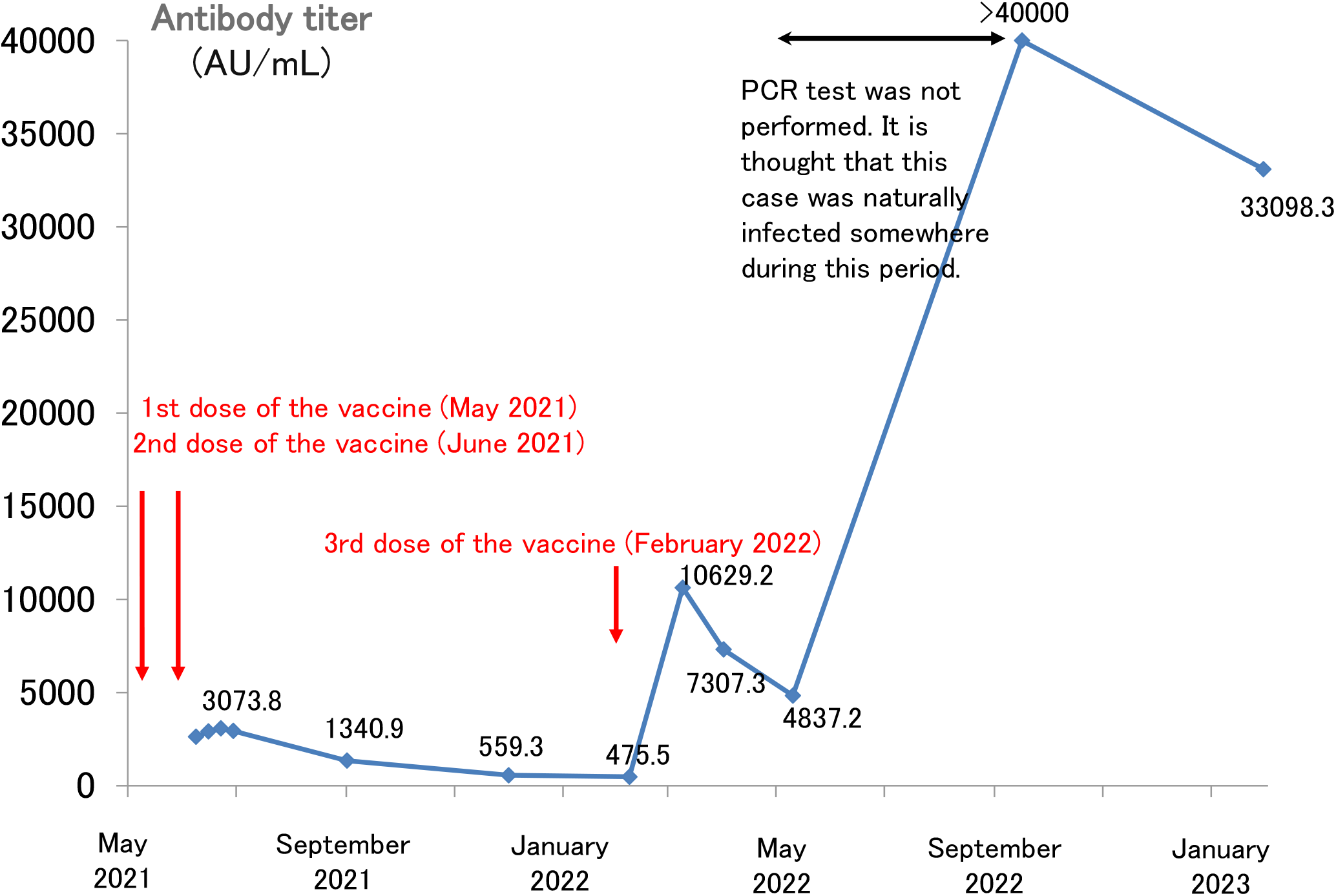
**A**. Longitudinal change of the SARS-CoV-2 antibody titer of a 40s male. **B**. Longitudinal change of the SARS-CoV-2 antibody titer of a 60s male. **C**. Longitudinal change of the SARS-CoV-2 antibody titer of a 30s female. **D**. The SARS-CoV-2 antibody titer of a 60s male.

There have been other cases of spontaneous COVID–19 infection after a second dose of the vaccine. A 60s male received a second dose of vaccine in April 2021 (Figure 4B). Thereafter, his antibody titer declined, temporarily falling to 600 AU/mL. However, he was spontaneously infected with COVID-19 in February 2022, and his antibody titer increased to over 40,000 AU/mL. When we followed up on the antibody titer in January 2023, nearly one year after the COVID-19 infection, the antibody titer was still high (> 10,000 AU/mL).

A 30s female had completed her second dose of vaccine by May 2021, and her antibody titer had decreased to approximately 300 AU/mL before the third dose of vaccine in February 2022 (Figure 4C). The third dose of the vaccine in February boosted her antibody titer to > 20,000 AU/mL. Although her antibody titer was boosted by the third dose of the vaccine, she spontaneously developed COVID–19 in May. After the COVID–19 infection, her titer increased to over 40,000 AU/mL. Six months after COVID–19 infection, the antibody titer remained high at 37,000 AU/mL.

There was a case of spontaneous COVID–19 infection after a third dose of the vaccine. A 60s male had completed his second dose of vaccine by June 2021, and received a third dose of vaccine in February 2022 (Figure 4D). After the third dose of vaccine, his antibody titer rose to more than 10,000 AU/mL, and then decreased to about 4,800 AU/mL in approximately two months. When the antibody titers were measured in September 2022, his titers increased to over 40,000 AU/mL. Like the case of Figure 4A, it is thought that this participant was spontaneously infected with COVID-19 somewhere from May to September 2022, but PCR testing was not conducted, and the participant was not counted as a COVID-19 infected patient. Even six months after the suspected COVID-19 infection, the antibody titer remained above 3,0000 AU/mL.

There were 17 cases of spontaneous COVID-19 infections after receiving two or more doses of vaccines (Table 2). All patients were infected with COVID-19 after two or three doses of the vaccine, and all of them were either asymptomatic or showed very mild symptoms. Some of them were not counted as infected persons because PCR tests were not performed. Eight of them had antibody titers of more than 4,0000 AU/mL after spontaneous infection, and two of them maintained antibody titers in the 10,000 AU/mL range even after more than six months had passed since infection.

**Table 2.** Cases of spontaneous COVID-19 infection after 2 or more doses of vaccine.

| Case | 1st<br>vaccination<br>date | 2nd vaccination<br>date | 3rd<br>vaccination<br>date | Date of infection | Post-infection<br>antibody test date<br>① | Post-infection<br>antibody titer①<br>(AU/mL) | Post-infection<br>antibody test date<br>② | Post-infection<br>antibody titer②<br>(AU/mL) | Post-infection<br>antibody test date<br>③ | Post-infection<br>antibody titer③<br>(AU/mL) |
| --- | --- | --- | --- | --- | --- | --- | --- | --- | --- | --- |
| 30s M | April 2021 | May 2021 |  | January 2022※ | February 2022 | >40000.0 | August 2022 | 31409.9 | November 2022 | 22684.0 |
| 60s M | April 2021 | May 2021 | February 2022 | April 2022 | April 2022 | >40000.0 | October 2022 | 15769.9 | December 2022 | 12379.0 |
| 40s M | May 2021 | May 2021 |  | August 2022 | September 2022 | >40000.0 | December 2022 | 19533.0 |  |  |
| 60s M | July 2021 | Aug 2021 | April 2022 | August 2022# | November 2022 | >40000.0 | December 2022 | 39247.6 |  |  |
| 60s M | April 2021 | May 2021 | February 2022 | August 2022 | November 2022 | >40000.0 |  |  |  |  |
| 40s M | July 2021 | August 2021 | July 2022 | August 2022 | November 2022 | 17451.4 |  |  |  |  |
| 20s F | April 2021 | May 2021 |  | January 2022 | February 2022 | >40000.0 |  |  |  |  |
| 20s F | May 2021 | June 2021 | March 2022 | August 2022 | November 2022 | >40000.0 |  |  |  |  |
| 20s F | April 2021 | May 2021 | January 2022 | August 2022 | November 2022 | 36446.4 |  |  |  |  |
| 20s F | July 2021 | August 2021 | April 2022 | August 2022 | December 2022 | 36292.0 |  |  |  |  |
| 30s F | April 2021 | May 2021 |  | January 2022 | January 2022 | 18907.4 |  |  |  |  |
| 20s F | August 2021 | September 2021 | April 2022 | December 2022 | January 2023 | 45218.1 |  |  |  |  |
| 20s F | April 2021 | August 2021 | May 2022 | August 2022 | December 2022 | 37648.8 |  |  |  |  |
| 20s F | July 2021 | August 2021 | April 2022 | August 2022 | December 2022 | 27238.4 |  |  |  |  |
| 20s F | July 2021 | August 2021 | March 2022 | September 2022 | December 2022 | >80000.0 |  |  |  |  |
| 60s F | May 2021 | June 2021 | March 2022 | December 2022 | December 2022 | 31879.8 |  |  |  |  |
| 40s F | March 2021 | April 2021 | December 2021 | January 2023 | February 2023 | 27849.0 |  |  |  |  |

※ No fever, only mild cold symptoms, PCR test was not conducted.

## Discussion

The COVID-19 vaccine has been available in Japan since February 2021 for healthcare workers and since April 2021 for the elderly citizens. As of December 10, 2021, 77.3% of the total population had completed two vaccination doses. The third dose of vaccinations also began in December 2021 for healthcare workers, and in early 2022, vaccinations began in earnest for the general population, starting with the elderly individuals.

Despite the expansion of COVID–19 vaccination, the Omicron strain has spread tremendously since the beginning of 2022. As of February 2023, the cumulative number of people infected with COVID–19 in Japan exceeded 32 million, most of whom were infected after 2022, when the Omicron strain became the dominant strain.

The elevation in and duration of the SARS-CoV-2 neutralizing antibody titer is considered an important indicator to confirm the efficacy of the COVID–19 vaccine (5). In addition, anti-receptor-binding domain (RBD) antibody, an antibody against SARS-CoV-2 spike protein, is widely used in Japan because it correlates with the SARS-CoV-2 neutralizing antibody titer (6,7) and can be measured at general medical facilities.

Antibodies against the SARS-CoV-2 spike protein after COVID–19 vaccination have been reported to decay over several months (8,9). In this study, consistent with previous reports, we confirmed that the SARS-CoV-2 antibody titer decreases with time after the second dose of the vaccine if there is no spontaneous COVID–19 infection.

Anti-RBD antibody titers are known to increase markedly after the booster dose (third dose) of the vaccine (10). In this study, consistent with previous reports, we have confirmed that the third booster vaccination is effective in increasing the antibody titer, but the boosted antibody titer after the third vaccination also decreased over time. This tendency is consistent with previous reports (11). Antibody titers before the third dose of the vaccine tended to correlate with the boosted antibody titer after the third booster dose. This result is also consistent with previous reports (12).

It has long been known that people who were previously infected with COVID–19 have markedly increased antibody titers after vaccination compared to uninfected people (13–15). It has been suggested that the vaccination of previously infected patients with COVID–19 is questionable (16). Breakthrough infections of COVID–19 after vaccination have become a problem. In Japan, breakthrough infections frequently occur after two or more vaccine doses. It has been reported that breakthrough infection after vaccination markedly increases antibody titers (17). However, there are currently few reports of long-term follow-up of antibody titers increased by breakthrough infections.

In this study, we observed that when spontaneously infected with SARS-CoV-2 after the second dose of vaccine, antibody titers increased to more than 40,000 AU/mL and remained high for several months. Although antibody titers decay after vaccination, it has been reported that not only memory B cells but also cellular immunity, such as memory T cells persist for six months after vaccination (18). The decrease in antibody titer may not directly lead to a decrease in infection protection, but the antibody titer may be immediately boosted by spontaneous infection if there is a history of vaccination in the past due to immune memory.

SARS-CoV-2 antibody titers vary greatly from person to person and increase prominently after spontaneous infection. In Japan, it is thought that many people have antibody titers of tens of thousands of AU/mL after spontaneous COVID-19 infection (often very mild and asymptomatic) following vaccination.

In Japan, a large number of patients are infected with the Omicron strain in 2022 after receiving two doses of the COVID–19 vaccine in 2021. As observed in this study, such cases are likely to maintain high antibody titers for more than six months after infection; however, in reality, even those with sufficiently high antibody titers are likely to have received a third or subsequent additional vaccination in a very large number of cases. It is questionable whether individuals who were infected with COVID–19, and whose antibody titers rose above the measurable limit and remained high for more than six months after infection, as was the case in the present study, should be uniformly vaccinated.

In conclusion, the increase in antibody titers against SARS-CoV-2 and the duration for which it remains high are considered important indicators of the efficacy of novel coronavirus vaccines. It is desirable to measure antibody titers and prioritize those with low antibody titers for booster vaccination, rather than blindly recommending booster vaccination to the entire population. As this was a small single-center retrospective study, larger-scale longitudinal studies of antibody titers are warranted.

## Data Availability

All data produced in the present study are available upon reasonable request to the authors.

## Notes

### Competing Interest Statement

The authors have declared no competing interest.

### Funding Statement

This study did not receive any funding.

### Author Declarations

The study protocols were approved by the ethics committee of Osaka Dental University Hospital (2022-10)

### Summary of Updates

I re-uploaded.

